# Refugee Healthcare Resilience and Burdens: A 10-year Mixed-Methods Analysis of System Shocks in Canada

**DOI:** 10.1101/2024.12.06.24318519

**Authors:** Eric Norrie, Linda Holdbrook, Rabina Grewal, Rachel Talavlikar, Mohammad Yasir Essar, Tyler Williamson, Annalee Coakley, Kerry McBrien, Gabriel E. Fabreau

## Abstract

**Background:** System shocks, including sudden policy changes, refugee surges and pandemics, strain healthcare systems. These shocks compound existing vulnerabilities in refugee healthcare, limiting ability to provide patient care, but can also catalyze resilient adaptations. Investigating how local refugee health systems respond to shocks is critical to understanding resilience.

**Methods:** We conducted a sequential explanatory mixed-methods study (2011–2020) at a specialized refugee health centre in Alberta, Canada, investigating four health system shocks: IFHP Funding Cuts (2012), Syrian Surge (2015), Yazidi Resettlement (2017), and COVID-19 (2020). We analyzed patient sociodemographic characteristics, health center utilization, and healthcare provider supply, conducting interrupted time series analysis of mean monthly appointments (total, family physicians, specialists and multidisciplinary team) and rates of change. We adapted a Health System Resilience framework to thematically analyze interviews with centre leaders and integrated these findings with quantitative findings to assess resilience and operational burdens.

**Findings:** From 2011 to 2020, 10,661 refugees from 106 countries attended 107,642 appointments. Mean monthly appointments rose from 455 to 2,208 (3.9-fold, p<0.01). Monthly appointments increased between IFHP and Syrian periods (610.8 to 937.9, p<0.01), but not between Syrian Surge and Yazidi Resettlement (p=0.29). During COVID-19, mean appointments remained stable (1,412.4 to 1,414.0, p=0.11), but additional monthly appointments rose from 6.3 to 110.4 (17.5-fold, p<0.01). Over ten years, mean provider hours increased from 320 to 736 (2.3-fold), and from 59.5 to 871.4 (14.6-fold) for family physicians and multidisciplinary team members. Qualitative analysis revealed resilience capacities but highlighted costs such as burnout, vicarious trauma, and financial strain. Integration showed the centre developed resilience but experienced notable operational burden.

**Interpretation:** Over a decade, a specialized refugee health centre adapted to successive shocks, transforming into a beacon clinic. It demonstrated resilience through care expansion and innovation, but with notable costs, financially and to health worker wellbeing.

**Funding:** None

## Introduction

Global forced displacement has reached historic levels, with 117 million people displaced as of 2023, the highest on record.^1^ This increase, driven by conflicts, persecution, disasters, both natural and climate-related, marks the twelfth consecutive year of rising displacement.^1^ Refugees face unique health vulnerabilities, including exposure to violence, poverty, and limited healthcare access that can strain health systems that receive large refugee influxes or face rapid policy shifts.^1–2^ The resilience capacity of health systems to address refugees’ complex needs remains critically underexplored.

Blanchet et al. developed a health system resilience framework that defines resilience through four dimensions: (1) *Knowledge*, the system’s ability to gather and apply diverse information; (2) *Uncertainties*, readiness to anticipate and manage unexpected events; (3) *Interdependence*, effective coordination within and beyond the health sector; and (4) *Legitimacy*, the ability to uphold norms and gain community trust.^3^ These dimensions shape a health system’s absorptive, adaptive, and transformative resilience capacities enabling it to maintain performance levels— defined by quantity, quality, and equity—under similar or reduced resources, or to restructure and enhance performance.^3^ Health system shocks, such as sudden policy shifts, rapid refugee influxes, and pandemics expose vulnerabilities by generating sudden demands or straining resources.^4^ Systems lacking resilience and adequate resourcing risk disruption when exposed to shocks, with system performance levels falling, failing to recover afterwards, or collapsing entirely under such pressures^.5,6^

Canada is a global leader in refugee resettlement, leading in annual formal resettlements from 2018 to 2022.^1^ However, unlike countries with formalized national refugee health systems, Canada relies on a decentralized approach, with most care delivered by primary care providers.^7^ Across Canada, specialized refugee ‘beacon clinics’ developed organically and act as hubs of expertise, that provide culturally sensitive, interdisciplinary care tailored to refugees’ unique needs^.8,9^ These centre often integrate primary and specialty care, mental health, public health, and social services to efficiently manage recently arrived refugees’ healthcare needs within Canada’s healthcare system.^8,9^ Over the past decade these centre have faced multiple health system shocks including, refugee health policy changes, rapid refugee influxes, and COVID-19 pandemics. Despite their critical role, their resilience capacities and adaptations to successive system shocks remains poorly understood.

Between 2011 and 2020, Canada’s refugee health system faced four major shocks.^10–14^ In 2012, cuts to the Interim Federal Health Program (IFHP), a national insurance program for refugees and asylum claimants, left many without essential services, such as medications and primary healthcare, increasing healthcare access barriers and forcing provincial systems to absorb emergency care costs for untreated conditions.^10^ The IFHP was reinstated in time to support the *Syrian Resettlement Initiative*, when Canada rapidly resettled 40,000 refugees between November 2015 and January 2017, creating a surge that strained local systems.^11,12^ From February 2017 to February 2020 the *Survivors of Daesh* program resettled 1,300 Yazidi refugees, survivors of genocide and horrific violence to four major Canadian cities. They arrived with complex trauma and physical health conditions that further strained refugee health services.^13,15^ In March 2020 the COVID-19 pandemic disrupted global health systems, adding strain and exacerbating health disparities between resettled refugees and host populations.^14^

Subsequent humanitarian crises in Afghanistan (August 2021^)16,^ Ukraine (January 2022), Sudan (April 2023), Gaza (October 2023), and Lebanon (October 2024) illustrate the urgent need for resilient health systems responsive to sudden forced displacement and other shocks.^17^ Despite multiple shocks, the impact on beacon refugee health centres remains unexamined, particularly their resilience capacities, adaptive strategies and impacts on healthcare providers and staff. This study addresses this gap by investigating health centre utilization, provider supply, provider and staff experiences and adaptations at one of Canada’s largest and longest-operating specialized refugee health centres to understand its response to consecutive health system shocks.^18^

## Methods

### Study Design and Setting

We conducted a sequential explanatory mixed methods study to investigate changes in utilization, provider supply, and perceived impacts associated with health system shocks at a specialized ‘beacon’ refugee health centre in Calgary, Canada between January 1, 2011, and December 31, 2020. Our study comprised three phases: quantitative, qualitative, and integration. In the quantitative phase, we analyzed patient sociodemographic characteristics, tracked changes in centre utilization, and measured provider work hours supply, using the latter two as proxies for system performance.

Our qualitative phase comprised of semi-structured interviews with centre leadership, supported by quantitative data visualizations, to understand experiences during each shock period. We conducted deductive thematic analysis of qualitative data, adapting Blanchet et al.’s health system resilience framework to interpret low resilience themes as *vulnerabilities*.^3^

Finally, we used an integration matrix to integrate quantitative and qualitative findings, assessing convergence and divergence to enable a comprehensive interpretation of system performance outcomes and operational burdens. We defined system performance outcomes using quantitative metrics for utilization and supply, categorizing it into five levels after a shock period:

1. *Collapse*: Performance falls until system failure.
2. *Disruption*: Performance declines below baseline to new lower level without recovery.
3. *Absorptive*: Despite shock, performance is maintained at pre-shock levels after shock period ends.
4. *Adaptive*: Performance is maintained or temporarily improves using fewer or different resources.
5. *Transformative*: Performance is sustainably improved at higher level compared to pre-shock baseline.^3,9^

We assessed operational burdens by examining integrated findings for financial strain, material shortages, provider well-being, quality of care, and infrastructure limitations.

### Health System Shocks

Our primary exposures of interest compared a baseline period to four distinct shock periods: 1) baseline period (January 2011–June 2012); 2) the IFHP Cuts (July 2012–October 2015); 3) the Syrian Surge (November 2015 to January 2017); 4) Yazidi Resettlement (February 2017–February 2020); and 5) COVID-19 (March 2020–December 2020).

### Quantitative Phase

We extracted electronic medical record (EMR) data from all patients who attended one or more centre appointments including sociodemographic, utilization, biometric, diagnostic testing, treatment, and clinical data. Sociodemographic information included age, sex, and country of origin, aggregated into World Health Organization (WHO) regions (i.e., African, Eastern Mediterranean, Americas, European, South-East Asia, Western Pacific) to ensure confidentiality.^19^ We analyzed patient appointments by provider type—family physicians, specialist physicians, and multidisciplinary team (MDT) members—using EMR scheduling software. We defined specialist physicians as non-primary care providers (e.g., internal medicine, pediatrics, obstetrics, psychiatry, infectious diseases, and hepatology) and MDT members as nurses, pharmacists, dietitians, and psychologists.

### Statistical Model

We used interrupted time series analysis to examine associations between shocks and monthly appointment volumes. We assembled linear regression models for four provider types: all providers, family physicians, MDT members, and specialist physicians. Quantitative analyses were conducted using Stata (version 18.0).^20^

For each study period, we modeled the monthly number of appointments (*Y_i_*) as a function of the intercept (*β*_0*j*_) and slope (*β*_1*j*_) for the *j*-th period. The general model for time point *i* in period *j* is:

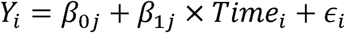

We used a linear combination of the intercept (*β*_0*j*_) and slope (*β*_1*j*_ ) coefficients to test for differences in the mean monthly appointments (level) and rate of change in appointments per month (slope) between adjacent periods, and overall change from baseline to COVID-19 period using a two-tailed alpha of <0.05. We generated data visualizations to facilitate qualitative interviews.

### Supply Analysis

We collected work hours for each family physician, specialist physician, and MDT member and categorized them by provider type. We calculated mean monthly averages and standard deviations for supply metrics for the baseline, each shock and over the entire study period.

### Qualitative Phase

We conducted semi-structured interviews with current and former centre leaders including medical directors, executive directors and managers to explore changes in utilization, adaptations and operational costs across shock periods. Interviews were conducted in person or online via Zoom between May and August 2022, based on participant preference.^21^ Participants were presented graphs summarizing quantitative data and prompted to reflect on utilization patterns, staff experiences, adaptive strategies and costs during each shock. Two researchers independently coded interview transcripts using deductive thematic analysis. We adapted Blanchet et al.’s framework to examine resilience dimensions—**knowledge** (the system’s ability to generate, integrate, and apply information effectively), **uncertainties** (the challenges related to unpredictable events), **interdependence** (the collaborative relationships that support system functionality), and **legitimacy** (the recognition and acceptance of the health center’s role in the community).^3^ We also examined **vulnerabilities** to investigate where resilience may have been lacking before the shock. Emerging themes were verified through iterative discussions and cross-checking to ensure consistency.

### Integration

We used an integration matrix to systematically combine and assess our data. Quantitative data provided system performance metrics, while qualitative insights captured resilience dimensions, including interdependencies, uncertainties, legitimacy, and knowledge, and pre-existing vulnerabilities. By analyzing convergence, divergence, and expansion across data types, we assessed each shock period for resilience, categorized as collapse, disruption, absorption, adaptation, or transformation. We also examined operational burdens incurred during each shock across five domains: financial strain, material shortages, provider wellbeing, quality of care, and infrastructure limitations.^22^

### Ethical Considerations

The University of Calgary’s Conjoined Health Research Ethics Board (CHREB – REB19-1029) approved the study and granted a waiver of consent for quantitative data. Informed verbal consent was obtained from all qualitative participants.

## Results

### Cohort Characteristics

We analyzed data from 10,661 patients representing 106 countries of origin (from all six WHO Regions^20^), who attended 107,642 appointments in total. The mean age was 24.5 years (SD 17.1), and 47.8% (5,094/10,661) were female. Table 1 summarizes sociodemographic characteristics by shock period and over 10 years.

**Table 1.** Cohort Characteristics by Time Period.

| Characteristic | Patients during<br>Baseline Period<br>Jan 2011 to May<br>2012<br>n=1710 (%) | Patients during<br>IFHP Cuts<br>Jun 2012 to Oct<br>2015<br>n=3391 (%) | Patients during<br>Syrian Surge<br>Nov 2015 to<br>Jan 2017<br>n=3191 (%) | Patients during<br>Yazidi Period<br>Feb 2017 to<br>Feb 2020<br>n=5377 (%) | Patients during<br>COVID-19 Period<br>Mar 2020 to Dec<br>2020<br>n= 1608 (%) | Patients during<br>All Time Periods<br>Jan 2011 to Dec<br>2020<br>n=10661 (%) |
| --- | --- | --- | --- | --- | --- | --- |
| Mean Years of Age at Intake<br>(SD)* | 26.5 (16.8) | 25.5 (16.7) | 25.0 (16.9) | 24.9 (17.5) | 24.3 (18.1) | 24.5 (17.1) |
| Sex |  |  |  |  |  |  |
| Female | 886 (51.8) | 1669 (49.2) | 1481 (46.4) | 2538 (47.2) | 794 (49.4) | 5094 (47.8) |
| Male | 824 (48.2) | 1722 (50.8) | 1709 (53.6) | 2832 (52.7) | 814 (50.6) | 5567 (52.2) |
| WHO Region of Origin |  |  |  |  |  |  |
| African Region | 684 (40.0) | 1626 (48.0) | 1321 (41.4) | 2287 (42.5) | 586 (36.4) | 4408 (41.3) |
| Eastern Mediterranean | 447 (26.14) | 823 (24.3) | 1253 (39.3) | 1868 (34.7) | 676 (42.0) | 3550 (33.3) |
| European Region | 55 (3.2) | 86 (2.5) | 30 (0.9) | 196 (3.7) | 12 (0.8) | 312 (2.9) |
| Americas | 163 (9.5) | 315 (9.3) | 262 (8.2) | 673 (12.5) | 227 (14.1) | 1169 (11.0) |
| South-East Asia | 168 (9.8) | 333 (9.8) | 180 (5.6) | 129 (2.4) | 25 (1.6) | 532 (5.0) |
| Western Pacific | 14 (0.8) | 50 (1.5) | 51 (1.6) | 52 (1.0) | 9 (0.6) | 103 (1.0) |
| Missing | 179 (10.5) | 158 (4.7) | 94 (3.0) | 172 (3.2) | 73 (4.5) | 595 (5.6) |
| Refugee Category |  |  |  |  |  |  |
| Government Assisted Refugee | 248 (14.5) | 1154 (34.0) | 1416 (44.4) | 2153 (40.0) | 940 (58.5) | 3991 (37.4) |
| Privately Sponsored Refugee | 224 (13.1) | 1173 (34.6) | 1065 (33.4) | 1456 (27.1) | 211 (13.1) | 2891 (27.1) |
| Asylum Claimant | 99 (5.8) | 311 (9.2) | 426 (13.5) | 1377 (25.6) | 330 (20.5) | 1815 (17.0) |
| Newborn Canadian | 28 (1.6) | 167 (5.0) | 114 (3.6) | 267 (5.0) | 107 (6.7) | 496 (4.7) |
| Missing | 1111 (65.0) | 586 (17.3) | 170 (5.3) | 124 (2.3) | 20 (1.2) | 1476 (13.8) |

\*SD denotes standard deviation from the mean.

**Table 2.**
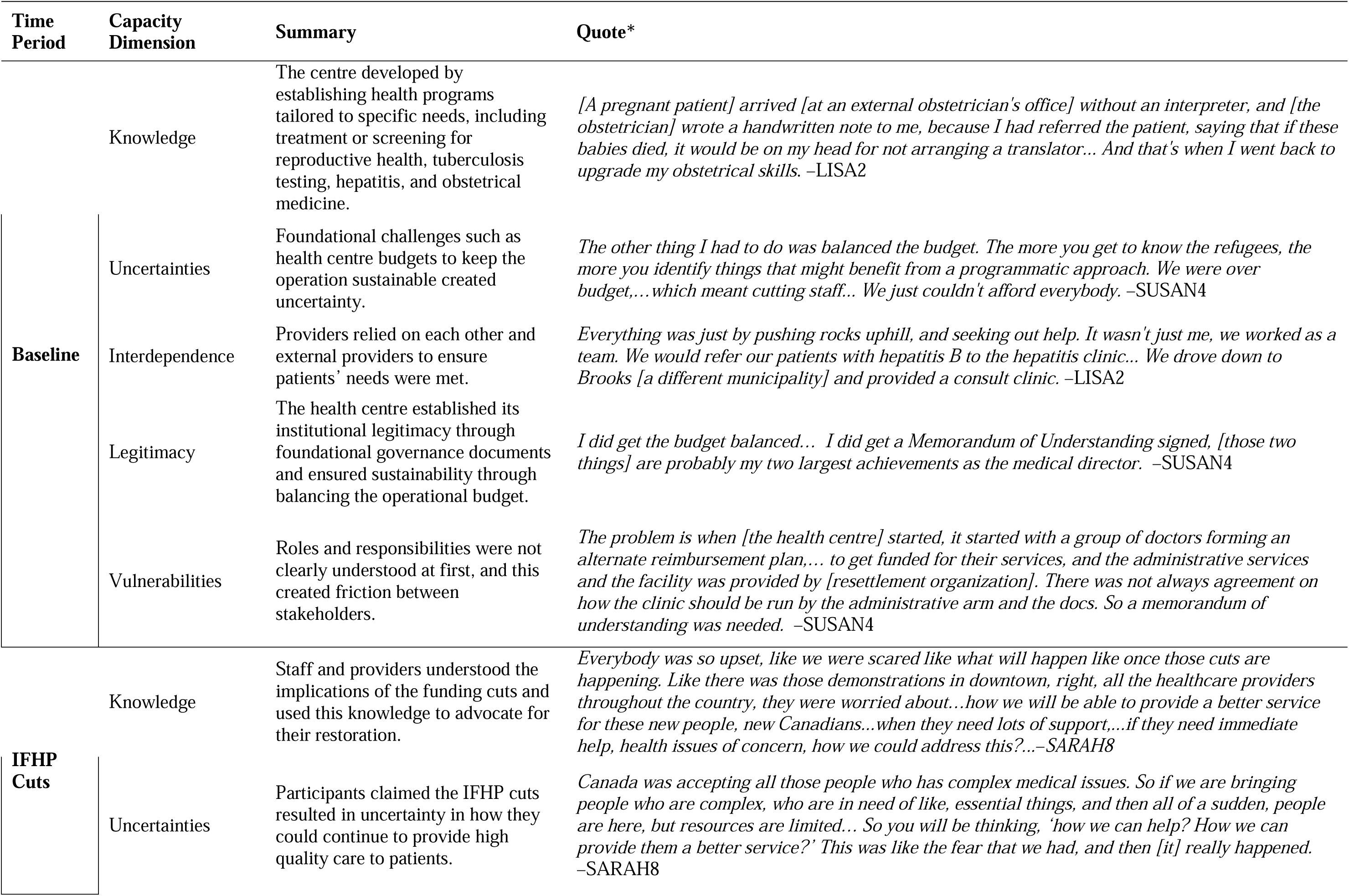
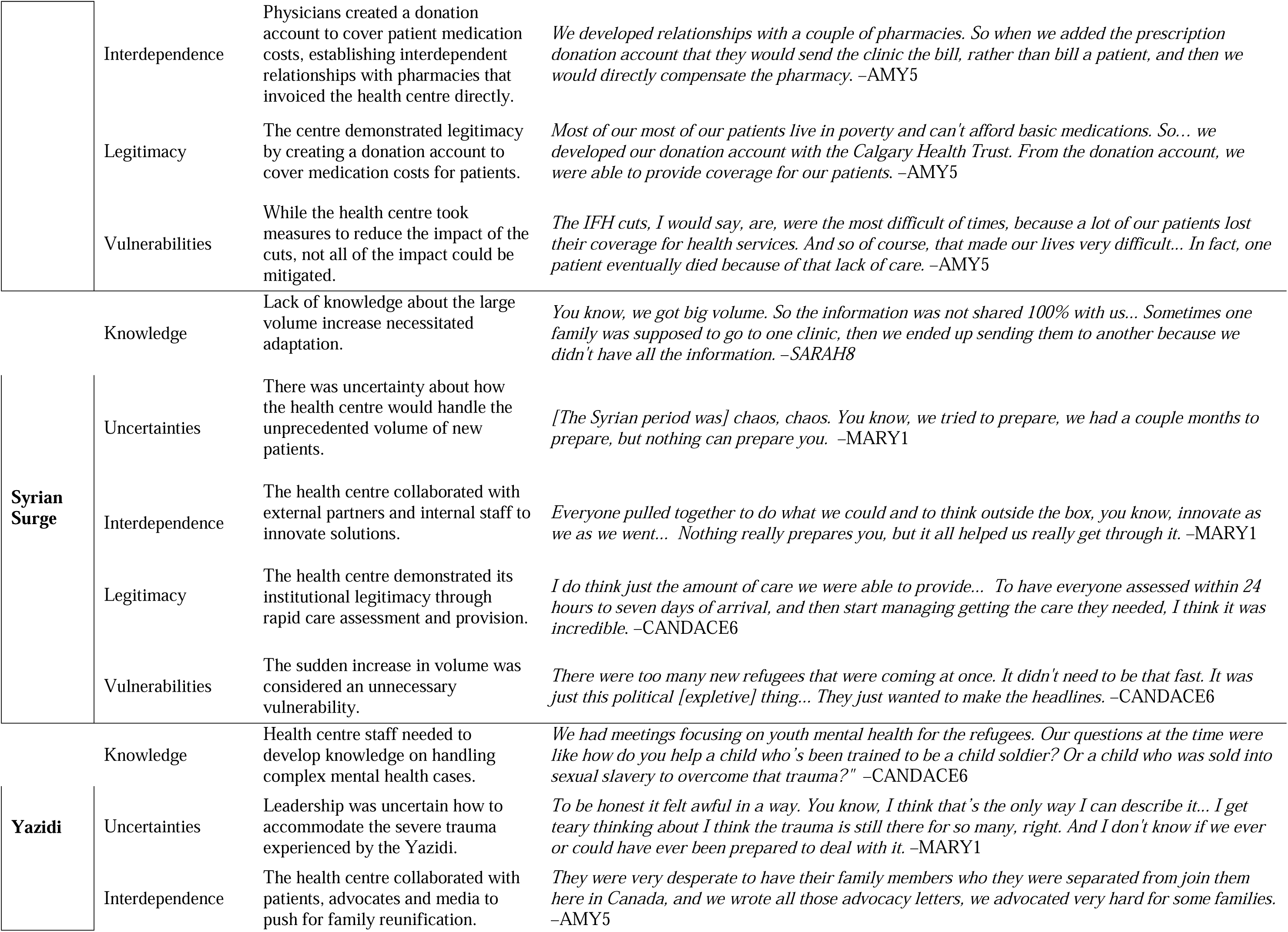
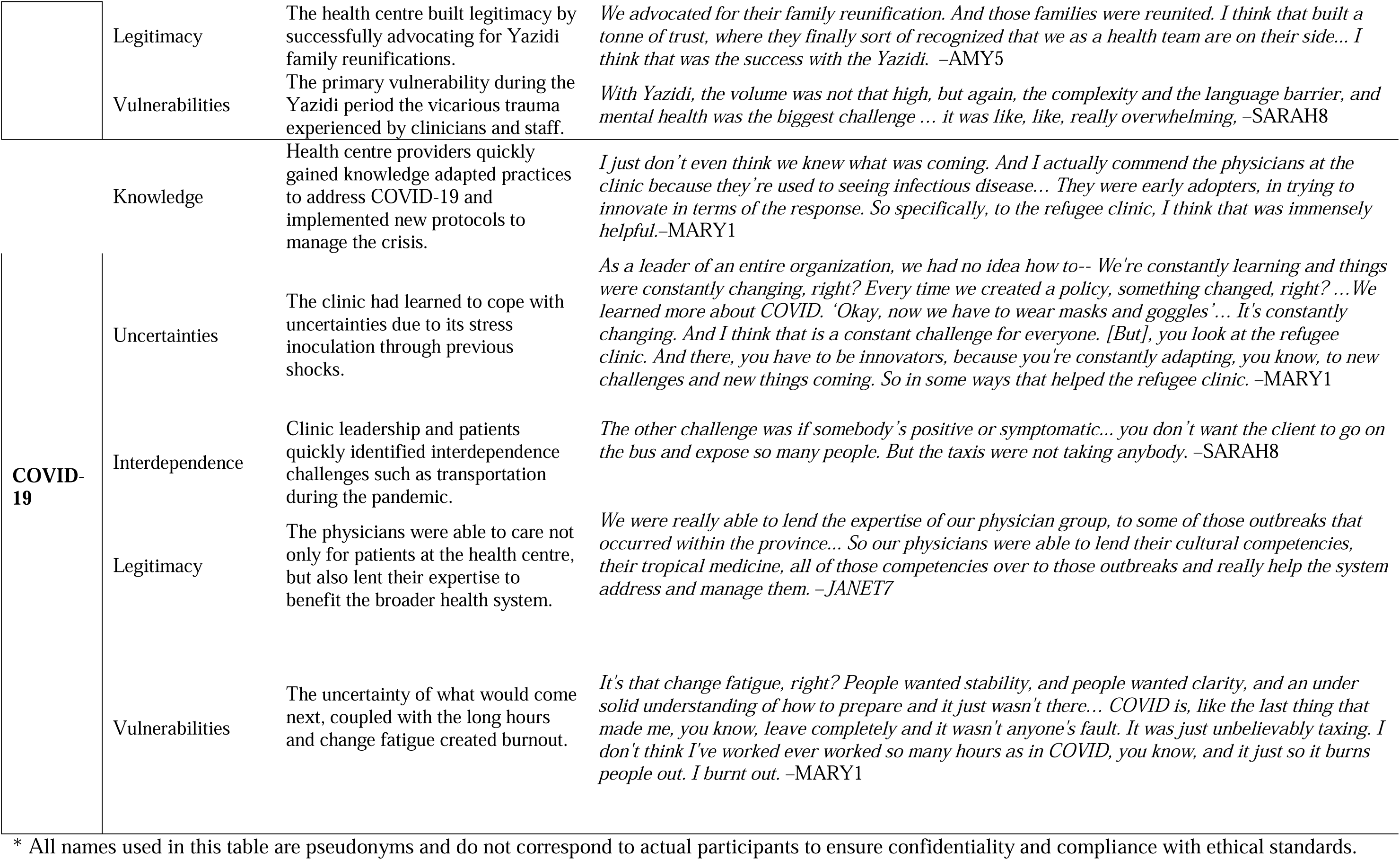
Health System Capacities.

**Table 3.** Integration Matrix.

| Shock | Quantitative Findings | Qualitative Findings* | Integrated Findings | Outcomes |
| --- | --- | --- | --- | --- |
| IFHP Cuts | Monthly family physician appointments remained stable, from 399 to 376.5 (p = 0.21). | *“The IFH cuts didn't really have an impact on our attended clinic appointments. It didn't really affect the number of patients that we saw, because we would see people whether or not they were covered by IFH or [provincial health coverage]. So we saw people, irrespective of their coverage... In fact, [we saw] new refugee claimants who had no coverage at all.” - AMY5* | **Convergence:** Quantitative stability aligns with qualitative findings of no major impact on clinic operations despite policy changes. | **Absorptive Capacity:** Quantitative and qualitative findings indicate that care levels mostly remained stable despite the shock. This was the most convergent capacity between our quantitative and qualitative findings. |
|  | Family physician supply remained similar, from 320 to 332 hours per month. | *“The IFHP cuts didn't actually impact us directly for the clinic operations. They impacted the refugees.” - SUSAN4* | **Expansion:** Qualitative data indicates that although utilization and provider supply were not impacted, it was a difficult time due to the increased burden on patients. | **Adaptive Capacity:** Clinicians adapted to the reduced financial resources by creating a donation account for patients to receive similar levels of care. This adaptation was temporary, not long term-transformation.<br>**Operational Burden:** The burden of this shock was primarily the **financial** (to patients directly, and to providers who donated to the trust account), and **quality of care**, as some patients were no longer able to get necessary medications, despite the providers’ efforts. |
| **Syrian Surge** | Significant increase in total appointments per month, from 610.8 to 937.9 ($p < 0.01$).<br>Family physician supply increased from 423.1 to 683.2 hours per month.<br>MDT supply increased from 152.8 to 366 hours. | *"I think what was beautiful was that the passion to serve was there. No matter how tired people were, they were still so invested in the work. You could just see it in the way people interact with the patients that would come into the clinic. I think that was really beautiful."* - CANDACE6<br>*"Everybody came together to discuss issues and meet the needs of the Syrians. It was a big community project, and everybody was enthusiastic and engaged... I think that those collaborations have served us well over the subsequent years."* - AMY5 | **Convergence:** Quantitative increases in appointments and provider hours are supported by qualitative reflections on intense collaboration and engagement.<br>**Divergence:** Qualitative accounts highlight the emotional investment and fatigue that quantitative data alone do not capture. | **Transformative Capacity:** Significant increases in all provider appointment change rate and doubling of MDT appointments.<br>**Operational Burden:** Heavy burden on **infrastructure** due to the sudden increase in volume but not a proportional increase in supply. Leadership perceived this "chaos" as impacting **provider wellbeing** by creating uncertainty and stress. |
| **Yazidi Resettlement** | No significant change in mean monthly appointments in any provider category. (All providers $p=0.29$, family physicians $p=0.86$, MDT $p=11$, specialists $p=0.21$)<br>No significant change in monthly appointment rate of change for any providers. (All providers $p=0.78$, family physicians $p=0.29$, MDT $p=0.10$, specialists $p=0.21$) | *"I am surprised that the numbers did not go up when you got the Survivors of Daesh. Certainly, they created a great deal more work for us. But we may not have had the psych support, because we didn't increase our psych support exponentially."* - SUSAN4<br>*"I think the beauty of the clinic is that we are adaptable... What really cinched it was when we did the advocacy for [the Yazidi]. They were very desperate to have their family members who they were separated from join them here in Canada, and we wrote all those advocacy letters, we advocated very hard... I think we did well in being flexible, adaptable, and being allies of our patients and going outside the box in terms of allyship."* - AMY5 | **Divergence:** Qualitative findings repeatedly emphasize the large perceived impact on staff and provider wellbeing which is not captured in the quantitative data.<br>**Expansion:** In addition to emphasizing the vicarious trauma experienced by staff and providers, lack of sufficient mental health support could explain the absence of quantitative care level increases. | **Adaptive Capacity:** The health centre advocated for family reunification to improve quality of care for patients.<br>**Absorptive Capacity:** No significant change in mean level or rate of change of overall monthly appointments, despite reported increased patient need.<br>**Operational Burden:** Large perceived impact on **provider wellbeing**. No significant change in clinic utilization and qualitative findings indicate that there lacked a proportionate investment to meet the increased mental health needs of patients, shifting some operational to quality of care. |
| COVID-19 | Significant increase in the slope of monthly appointments, from 6.3 to 110.4 additional appointments per month ($p < 0.01$).<br>Specialist monthly supply hours were nearly halved, from 78.6 hours per month to 41.9 hours per month). | *We were so busy with the other stuff, dealing with all these pandemic problems like COVID... We were busy, but we changed the workstyle. So like, from in-person, to virtual appointments, lots and lots of phone appointments. And then we were part of all these COVID vaccine clinics. -SARAH8*<br>*It's our same panel, but more frequent follow up. And I think that's because of the whole issue with telephone appointments and working through language barriers and cultural barriers and health literacy barriers... Also, our patients are at higher risk of complications of COVID-19, it would have required more frequent follow up. So I think that's what's responsible for that uptick. -AMY5* | **Convergence:** Quantitative data on rising appointments and provider hours aligns with qualitative accounts of constant adaptation and innovative responses to the pandemic.<br>**Expansion:** The significant drop in specialist appointments is the only sustained decrease in a quantitative metric in our visualizations. | **Adaptive/Transformative Capacity:** Highly significant increases in appointment rate of change. Virtual appointments demonstrate an adaptation to the pandemic. If sustained this would indicate a transformation.<br>**Disruption of Specialist Provider Services:** Monthly specialist provider hours per month fell from 78.6 to 41.9, close to the baseline level of 37.2 hours per month.<br>**Operational Burden:** Qualitative participants described staff and provider burnout and reduced **wellbeing**. Perceived **patient care** was reduced due to technology and language barriers. |
| Overall Resilience | Overall increase from first to last study time point: from 455 to 2208 monthly appointments ($p < 0.01$).<br>Family physician supply increased more than twofold; MDT supply increased over 14-fold. | *"We started with two rooms, and [look] where it is today. It is astonishing to see what made the difference every step of the way." - LISA2*<br>*"We are very flexible. The team would give you support material, respecting client's tradition, culture, [instructing] how to adapt, how to be open, how we could welcome them, by making things flexible." - SARAH8* | **Convergence:** Qualitative findings corroborate the large increases in clinic utilization and provider supply.<br>**Expansion:** Qualitative findings also indicate the health centre aimed to be flexible and meet patients' needs for cultural sensitivity. | **Transformative Capacity:** Large, sustained improvements across care utilization, provider monthly hours, and perceived quality of care.<br>**Operational Burden:** To accomplish this transformation over ten years the health centre expanded its **infrastructure, workforce, materials** such as interpretation services and protective masks, and **finances** to pay for these improvements. Responses to shocks often came at the expense of clinician **wellbeing**. |

\*All names used in this table are pseudonyms and do not correspond to actual participants to ensure confidentiality and compliance with ethical standards.

### Interrupted Time Series Analyses

#### All Providers

Data from January–February 2011 were incomplete and excluded, leaving 118 months for analysis (Figure 2). Overall, mean monthly appointments increased 4.9-fold, from 455 at baseline to 2,208 during the COVID-19 period (p<0.01). The rate of change for monthly appointment rose 69-fold, from 1.6 to 110.4 additional appointments per month (p<0.01) (Figure 1). Between the baseline to the IFHP Cuts period, neither mean monthly appointments, nor the rate of change of additional monthly appointments, changed significantly (p=0.16 and p=0.45 respectively). However, between the IFHP Cuts and Syrian Surge periods, mean monthly appointments increased from 610.8 to 937.9 (p< 0.01), though the rate of change did not differ, (4.5 to 9.1 additional appointments per month; p=0.48). Neither the mean appointments (1073.9 to 1179.9; p=0.29) nor the rate of change (9.1 to 6.3 additional appointments per month; p=0.78) differed between the Syrian Surge and Yazidi Resettlement periods. Between the Yazidi and COVID-19 periods, mean monthly appointments did not change (1413.9 to 1414.4, p=0.11), but the rate of appointments increases rose dramatically 17.5-fold, from 6.3 to 110.4 additional appointments per month (p<0.01).

**Figure 1.**
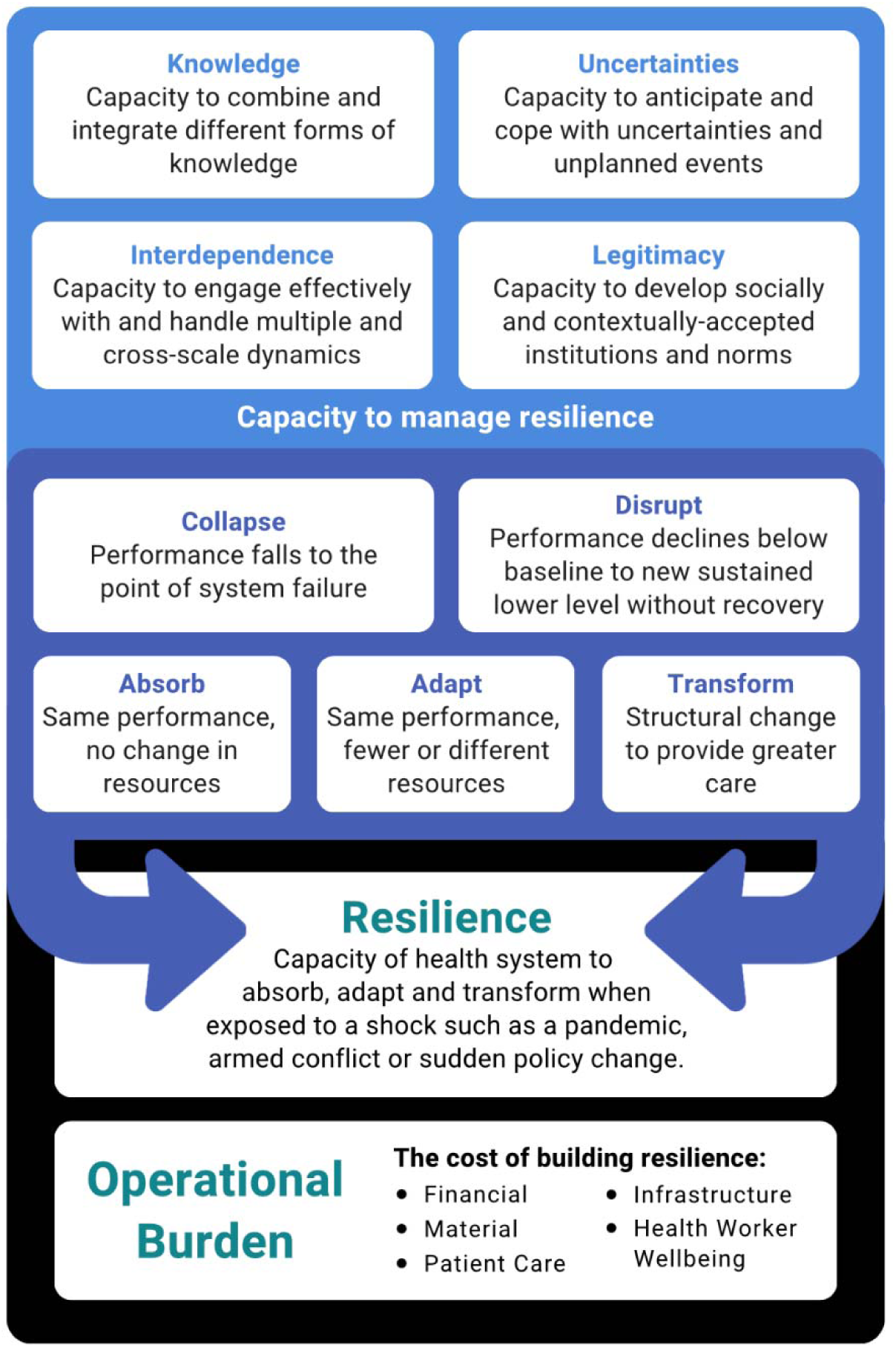
Resilience and Burden Framework, adapted from Blanchet, et al. (2017)

**Figure 2.**
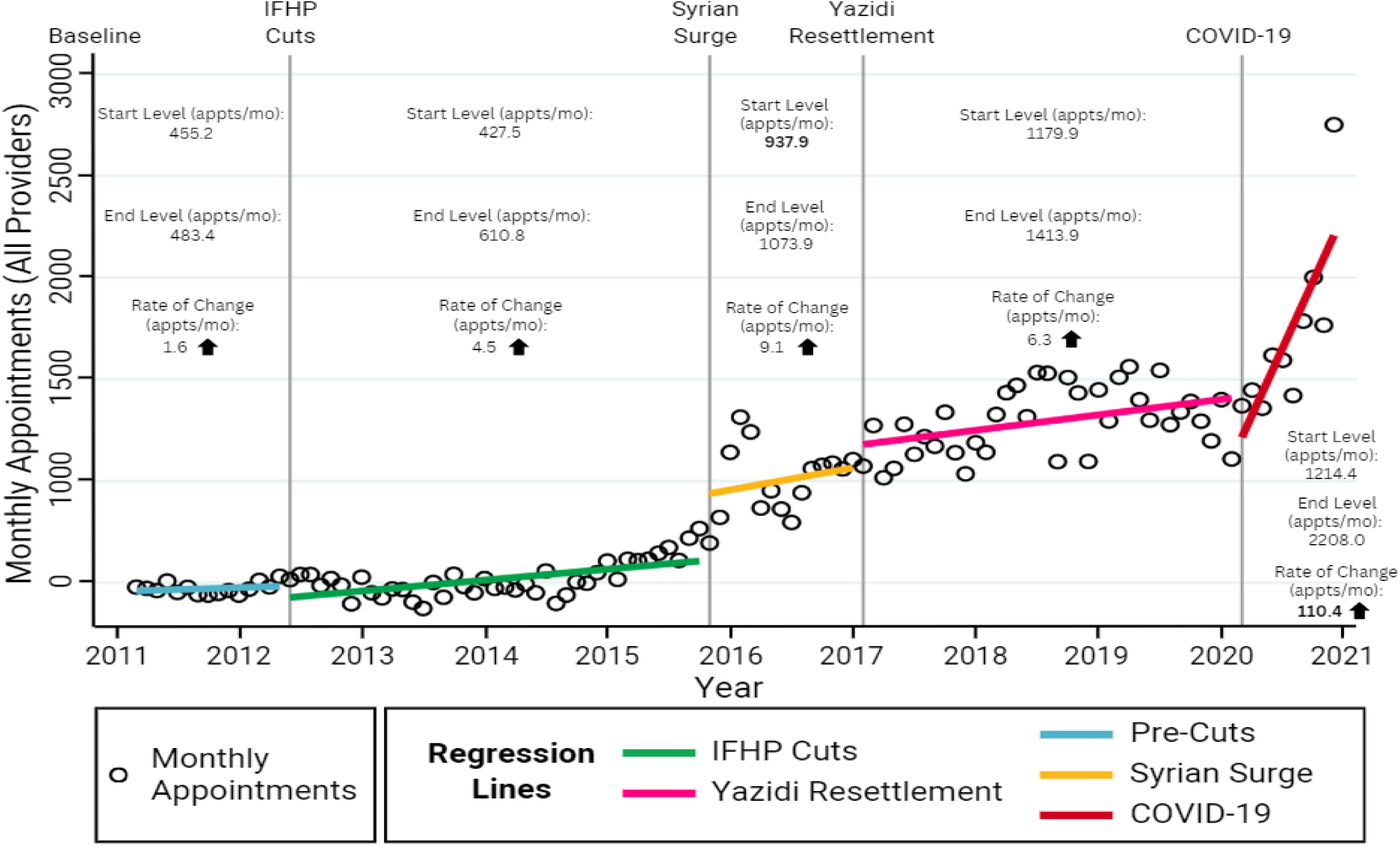
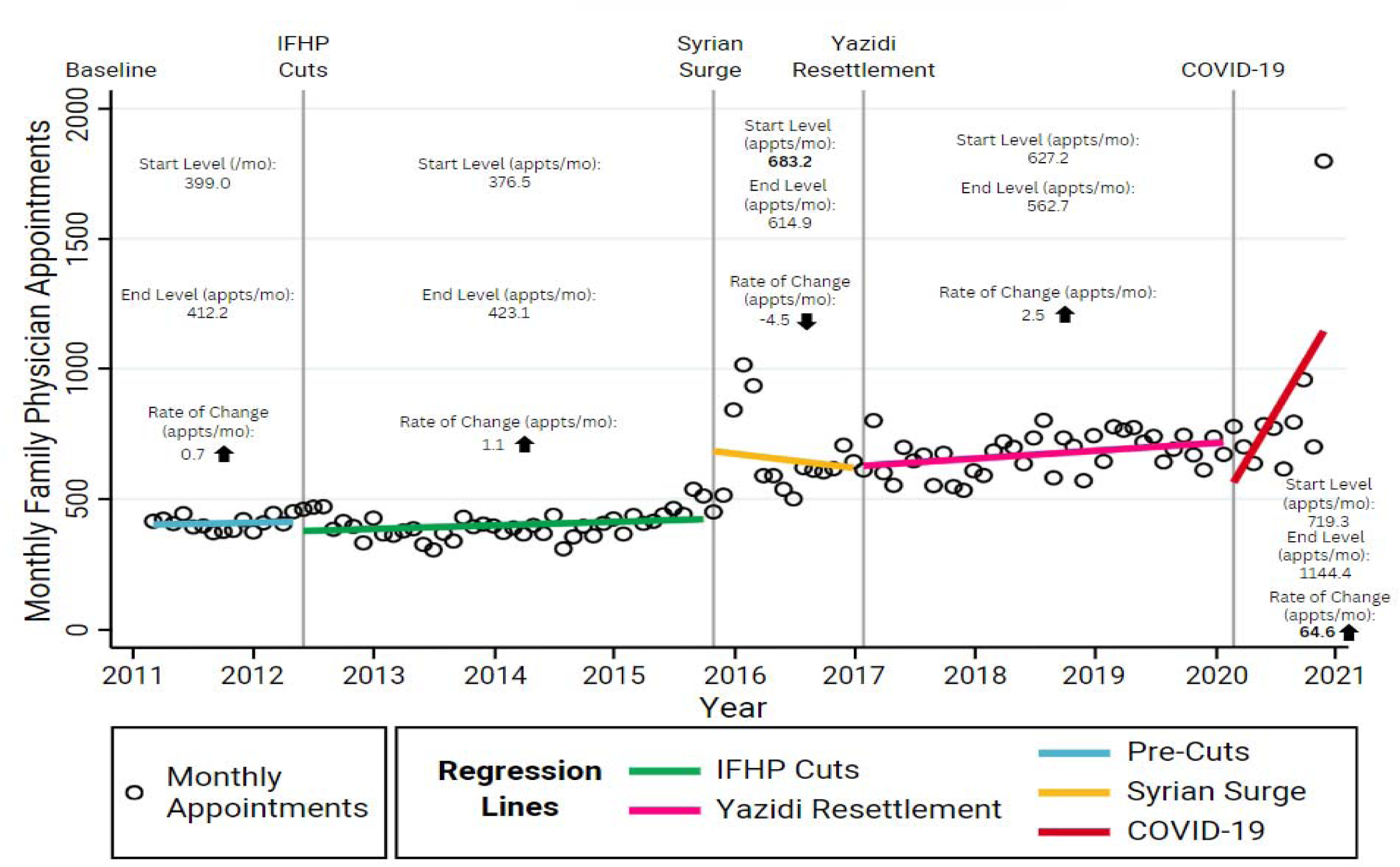
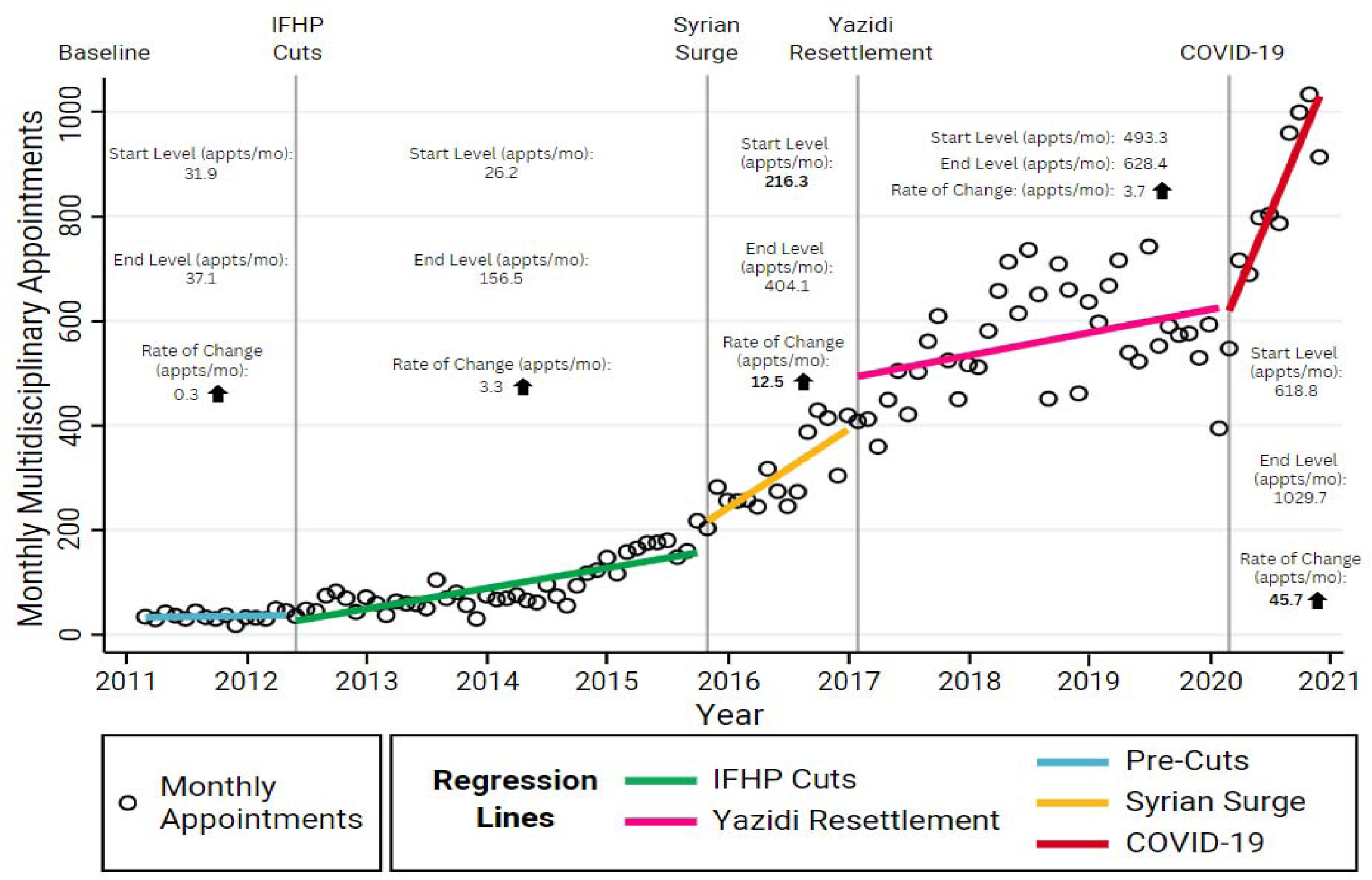
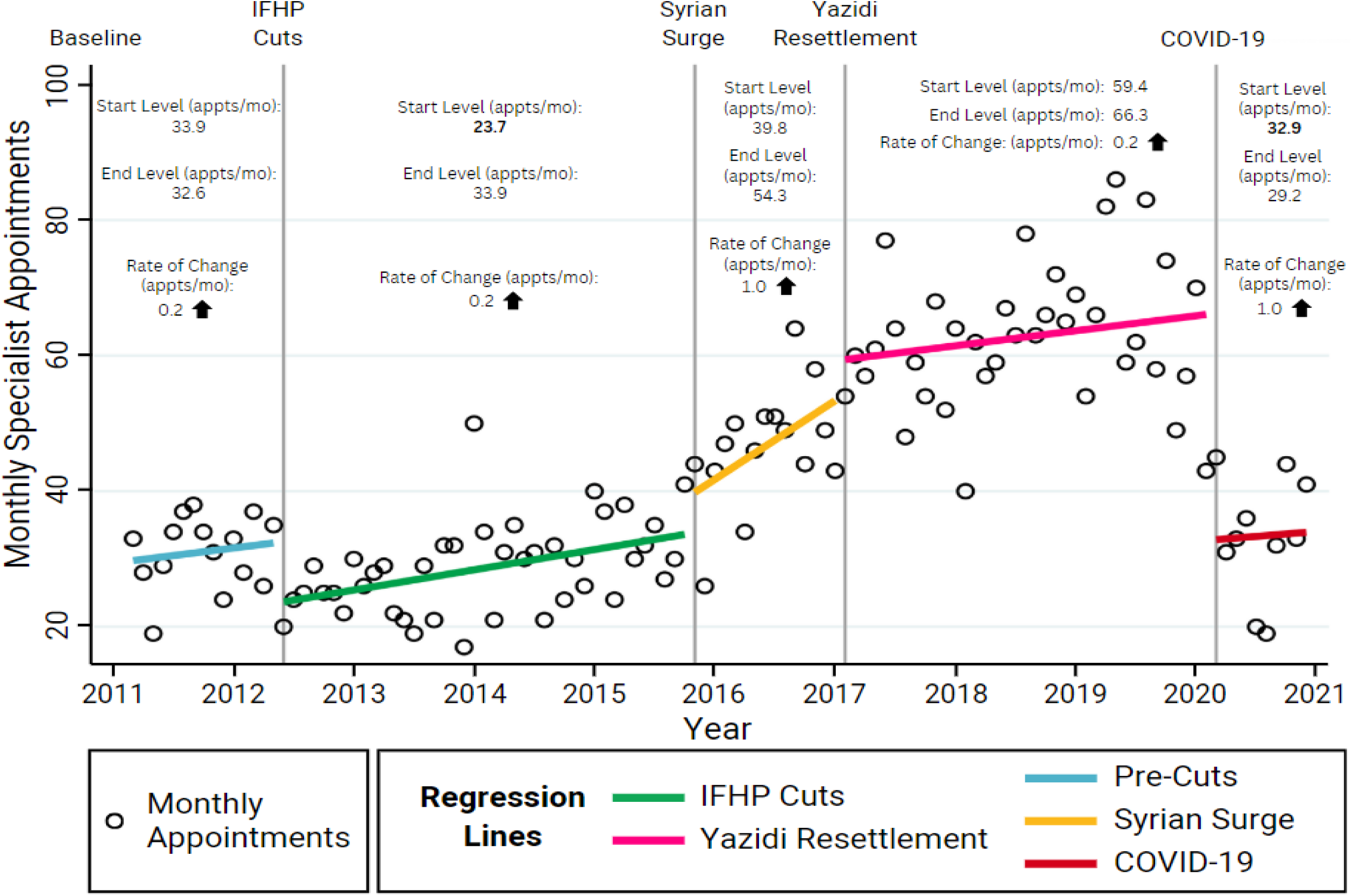
Monthly Appointments Across Shock Periods by Provider Type. **a. All Providers:** Monthly appointments for all providers are displayed from March 2011 to December 2020. A significant increase in appointment levels occurred between the IFHP Cuts and Syrian Surge periods (427.5 to 610.8, p < 0.01). During the COVID-19 period, the rate of monthly appointment changes increased dramatically (6.3 to 110.4 appointments/month, p < 0.01). **b. Family Physicians:** Monthly appointments with family physicians are shown between March 2011 and December 2020. Appointment levels increased significantly from the IFHP Cuts to Syrian Surge periods (376.5 to 683.2, p < 0.01). A substantial increase in the rate of monthly changes was observed during COVID-19 (64.6 appointments/month, p < 0.01). **c. Multidisciplinary Team (MDT):** Monthly MDT appointments are displayed from March 2011 to December 2020. Significant increases were observed in both levels and rates of change, including a level increase from the IFHP Cuts Final (26.2) to Syrian Surge Final (156.5, p = 0.01), and a rate increase from Yazidi (3.7) to COVID-19 (45.7, p < 0.01). **d. Specialist Physicians:** Monthly specialist physician appointments are shown between March 2011 and December 2020. A significant decrease in appointment levels was observed during COVID-19, dropping from Yazidi Resettlement Final (66.3) to COVID-19 Initial (32.9, p < 0.01). No significant changes in the rate of monthly appointments were observed across the shock periods. **Annotations:** Start and end levels (appointments/month) and rates of change are labeled for each period. Bolded values denote statistical significance (p < 0.05) at a two-tailed alpha level of 0.05.

**Figure 3.**
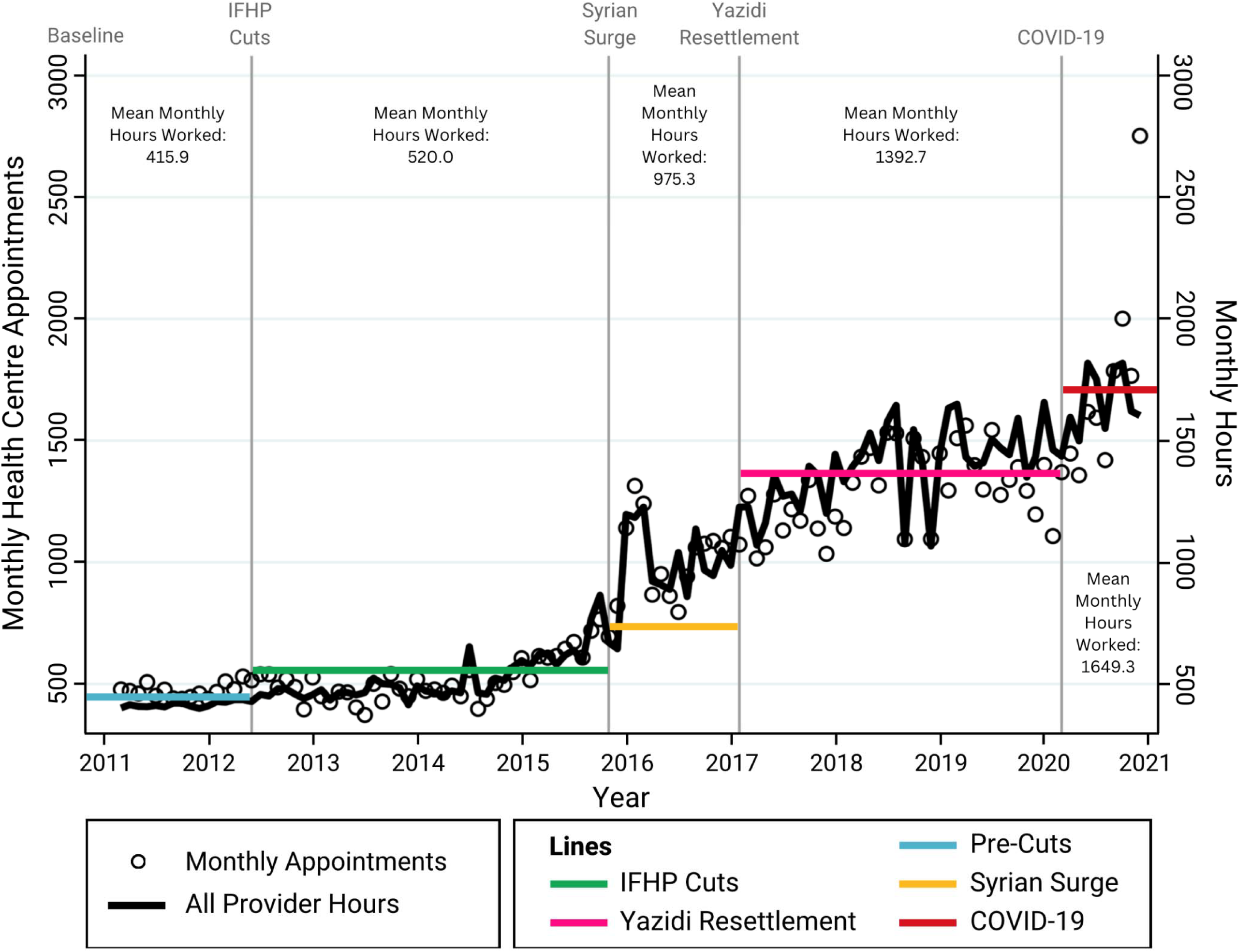
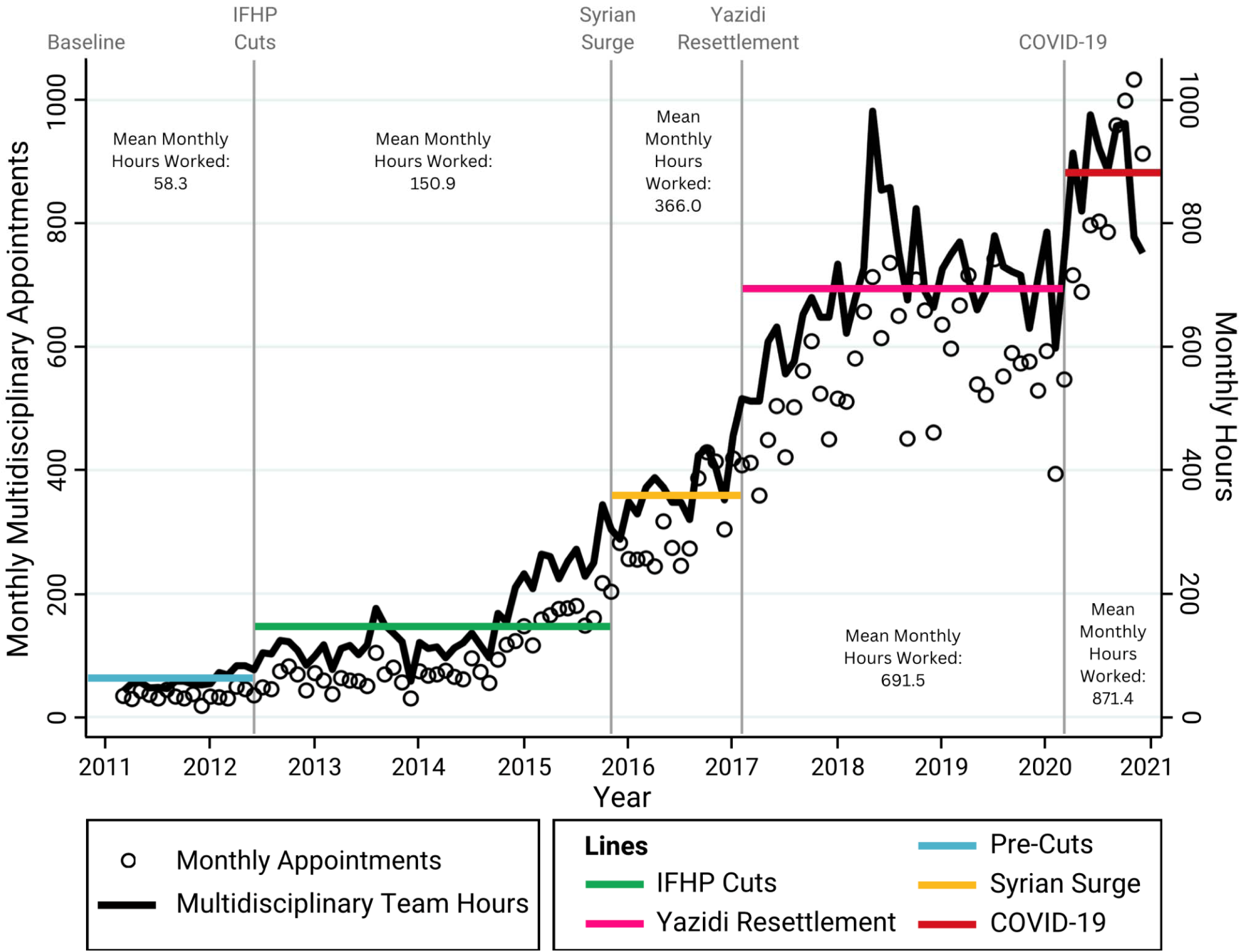
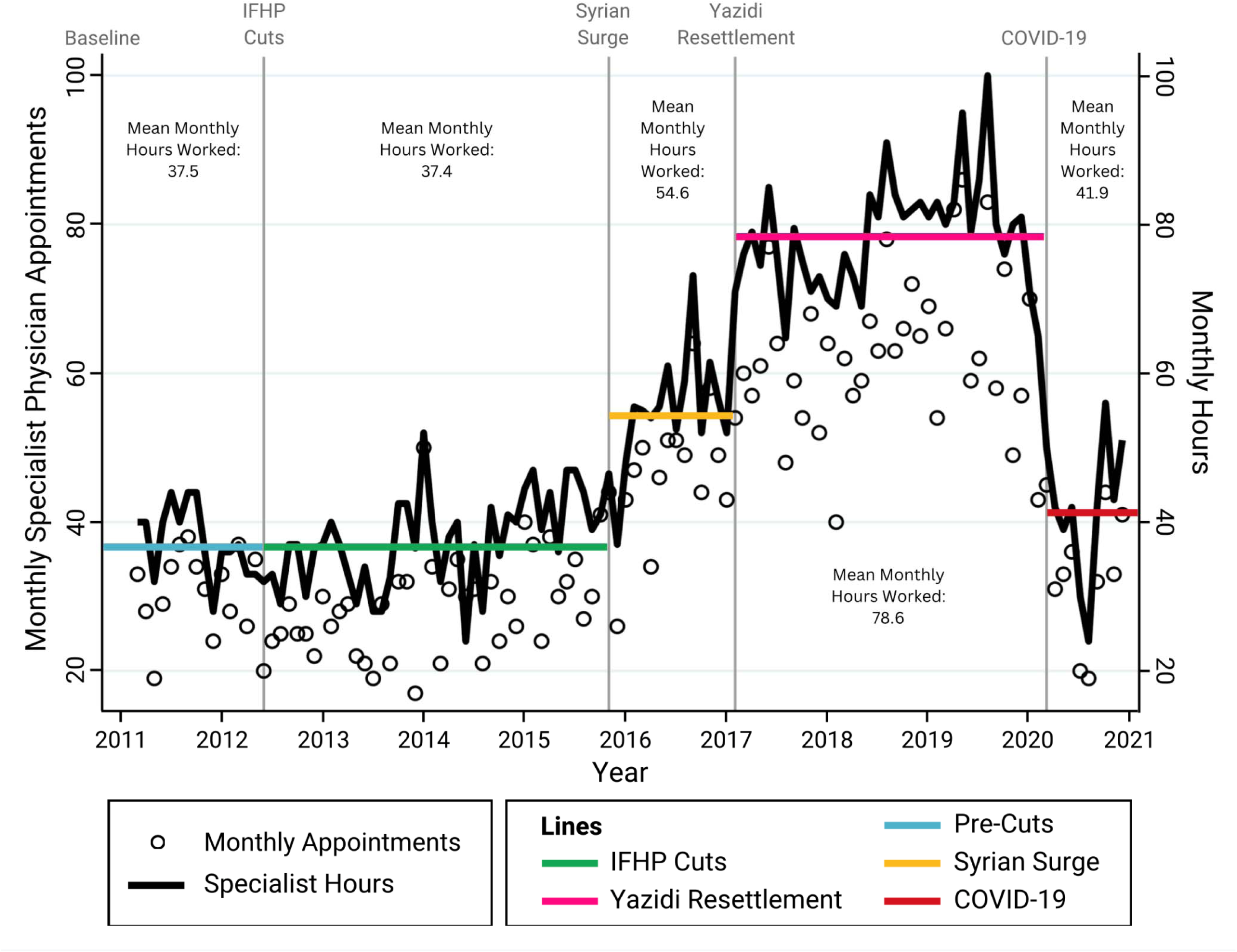
Provider Hours and Appointments Across Shock Periods. **a. Overall Provider Supply and Patient Utilization:** Monthly provider hours (black line) are displayed alongside monthly appointments (black circles) for all providers between March 2011 and December 2020. Horizontal lines indicate the mean provider hours worked during each shock period: Baseline, IFHP Cuts, Syrian Surge, Yazidi Resettlement, and COVID-19. **b. Family Physician Supply and Patient Utilization**: Monthly family physician hours (black line, scaled 5:1) are overlaid on monthly family physician appointments (black circles) between March 2011 and December 2020. Horizontal lines show the mean hours worked by family physicians during each shock period. **c. Multidisciplinary Team (MDT) Supply and Patient Utilization:** Monthly MDT hours (black line) are displayed alongside monthly MDT appointments (black circles) from March 2011 to December 2020. Horizontal lines represent the mean hours worked by MDT members during each shock period. **d. Specialist Physician Supply and Patient Utilization:** Monthly specialist physician hours (black line) are overlaid on monthly specialist appointments (black circles) between March 2011 and December 2020. Horizontal lines illustrate the mean hours worked by specialist physicians for each shock period.

### Appointments by Provider Type

Figure 2b illustrates the increase in family physician appointments, with mean monthly appointments rising 2.9-fold, from 399 at baseline to 1,144.4 during COVID-19, and the rate of monthly changes increasing 87.3-fold, from 0.7 to 64.6 additional appointments per month (both p<0.01). MDT appointments also rose dramatically, increasing 32.3-fold in mean monthly appointments, from 31.9 at baseline to 1,029.7 during COVID-19, with the rate rising 157.6-fold, from 0.29 to 45.7 additional appointments per month (both p<0.01) (Figure 2c). Specialist appointments, by contrast, did not differ in the rate of change of monthly appointments (Figure 2d). Mean monthly specialist appointments rose modestly from 33.9 at baseline to 39.8 during the IFHP Cuts period (p=0.02) but declined significantly during COVID-19, dropping from 66.3 to 32.9 (p<0.01). Consequently, the mean monthly specialist appointments at baseline (33.9) did not differ from the final level during COVID-19 (29.2; p=0.46).

### Provider Supply

Provider work hours increased significantly for family physicians and MDT members, reflecting supply growth to accommodate rising centre utilization. Family physician hours rose from 320 hours per month at baseline to 736 hours per month during the COVID-19 period, (2.3-fold increase). MDT supply increased 14.9-fold, expanding from 58.3 hours to 871.4 hours per month. In contrast, specialist physician supply initially rose from 37.5 hours at baseline to 78.6 hours during the Yazidi Resettlement period, but declined nearly 50% during COVID-19, dropping to 41.9 hours per month (eTable 3).

### Qualitative Results

We interviewed eight past and present refugee health centre leaders including executive officers, medical directors, and managers between May and August 2022. The interviews lasted between 30 to 60 minutes. Participants mostly identified as white women aged 35 to over 65 years. Table 5 presents themes and subthemes of resilience dimensions and vulnerabilities across all five study periods.

During the baseline and IFHP cuts periods integrating and applying different forms of knowledge was crucial, though finances emerged as a key vulnerability. During the Syrian surge period interdependence with health and resettlement partners was critical for handling the rapidly increased patient volume. In the Yazidi Resettlement period, staff and provider wellbeing emerged as a significant vulnerability due to vicarious trauma, while the centre developed legitimacy among the Yazidi community through advocacy for family reunification. During the COVID-19 pandemic, participants emphasized adaptability, building on lessons from previous shocks to navigate the uncertainties of COVID-19.

### Integration

The health centre demonstrated absorptive, adaptive, and transformative capacities, but these came with operational burdens. Over the 10-year study period, the health centre demonstrated transformative resilience, as evidenced by a 4.9-fold increase in mean monthly appointments from 455 at baseline to 2,208 by the COVID-19 period (p<0.01), and significant expansions in provider hours, with family physician hours more than doubling, and MDT hours increasing 14.6-fold (Figure 2). These dramatic expansions of system performance reflect the health center’s capacity to adapt and transform in response to recurring shocks.

During the IFHP Cuts period, care levels were maintained despite policy-driven financial strain on patients and moral distress among providers who worked and personally donated funds to fill care gaps, demonstrating absorptive capacity. The Syrian Surge marked a period of transformative capacity, with mean monthly appointments rising rapidly from 610.8 to 937.9 (p<0.01), supported by substantial increases in provider hours and collaborative adaptations among stakeholders. However, this surge also led to significant operational burdens, including increased provider burnout and infrastructure strain due to the high patient volume. During the Yazidi Resettlement, service levels were maintained despite high mental health needs that created additional burdens on staff, many of whom reported vicarious trauma. The centre adapted to these mental health needs through advocacy for family reunification which built community trust, and a vicarious trauma training program for staff and providers. The COVID-19 pandemic further illustrated the centre’s adaptability, as it rapidly increased overall care levels and provider supply. Yet, this period also saw a collapse in specialist care, with appointments halving from 66.3 to 32.9 per month and provider hours dropping from 78.6 to 54.6. While these adaptations highlighted the centre’s flexibility and resilience, they came with accumulated operational burden, including expanded infrastructure, increased human resources, financial costs, and challenges to provider wellbeing, such as burnout and change fatigue. Despite these challenges, the health center’s ability to navigate these shocks reflects its role as a model for transformative resilience in refugee health systems.

## Discussion

To our knowledge, this is the largest 10-year longitudinal cohort study of resettled refugees in a primary care setting in Canada, offering insights into how over a decade a specialized refugee health centre absorbed, adapted, and transformed in response to multiple health system shocks. Through a mixed-methods approach, we observed significant increases in utilization and provider supply, and identified resilience capacities—absorptive, adaptive, and transformative— alongside operational burdens during four shock periods. These findings reflect the centre’s operational resilience and the continuous demands imposed by external shocks.

Over the decade, both appointments and provider hours increased substantially, with family physician and multidisciplinary team appointments rising by 2.9-fold and 32.3-fold, respectively. This growth reflects the health centre’s transformation into a beacon clinic that expanded its services to meet patient needs through specialized programs while increasing overall capacity. This transformation enhanced refugee care and enabled the centre to contribute expertise within the province. During the COVID-19 pandemic the health centre demonstrated its strongest adaptation, as the rate of monthly appointments rose steeply (from 6.3 to 110.4 additional appointments per month). However, specialist care significantly declined, due to redeployment of hospital-based specialists to manage COVID-19 surges. This decline mirrored broader disruptions in Canadian healthcare, as surgeries and hospital admissions dropped significantly between March 2020 and June 2021, disproportionately affecting low-income individuals.^23^ Nonetheless, despite the collapse in specialist care, the health center’s resilience grew over time but was not an assured outcome of each shock.

Study data integration assessed resilience capacities and operational burdens across each shock period. The IFHP Cuts demonstrated strong absorptive capacity, as family physician appointments were maintained despite funding cuts; however, financial strain on patients and moral distress among providers, who donated funds to address gaps in medication coverage, revealed substantial operational burdens. In contrast, the Syrian Surge, demonstrated transformative capacity, with sustained increases in appointments and provider supply that exceeded pre-shock levels. This period also fostered sustained transformative collaborations and partnerships with settlement and other sectors to coordinate services. During the Yazidi Resettlement, the health center exhibited adaptive capacity by maintaining service levels and advocating for family reunification, despite high mental health needs that contributed to vicarious trauma among staff and providers.^24^ Finally, the initial months of COVID-19 revealed absorptive and adaptive capacities as the centre rapidly increased appointments and provider supply, leveraging virtual appointments, to address a sudden surge in their patients’ outpatient health needs. If sustained post-pandemic, these adaptations may represent transformative capacity.

The WHO Health System Building Blocks provide a foundational framework for understanding health system functions, but does not address how systems respond to shocks.^25^ Blanchet et al.’s resilience framework was instrumental to understand the refugee health center’s resilience capacities such as knowledge application, interdependence, uncertainty management, and legitimacy.^3^ Similarly, other studies have similarly applied this framework to examine the impacts of shocks like COVID-19 and the Syrian surge; ^26,27^ however, while it effectively addresses shock responses, the framework does not integrate structural elements such as the WHO Building Blocks, or outcomes of resilience processes. Other published frameworks integrate shock preparedness with elements of the Building Blocks, or include bottom-up perspectives on everyday resilience that highlight the critical role of frontline actors, but also fail to explicitly address resilience outcomes.^28,29^ Our study contributes to this evolving discussion by incorporating dimensions such as disruption, collapse, and operational burdens—including financial strain, material shortages, provider well-being, and infrastructure challenges.

By integrating qualitative and quantitative data, our findings demonstrated that resilience can be understood as both a process and an outcome. ^4,30^ While other studies caution against framing resilience as an outcome, our mixed-methods approach indicate that resilience indeed can be interpreted both as system processes and an outcomes.^4,30^ Processes were captured through our qualitative analysis revealing how frontline actors adapted during shocks, while outcomes were captured through quantitative analysis that showed sustained system performance despite shocks. Critically, our study highlights that system-level resilience likely stems from both systemic capacities and the adaptability of individuals within systems—whose stress inoculation responding to previous shocks likely contribute to system resilience. By incorporating frontline perspectives, we avoid misattributing resilience solely to the system, and instead recognize how individuals interact with their environment to build resilience over time.

Our findings align with the Johns Hopkins Center for Humanitarian Health – Lancet Commission’s call for critical research into resilient health systems capable of adapting to ongoing crises.^17^ Our decade-long analysis of over 10,000 refugee patients from 106 countries, contextualized by refugee health leaders’ firsthand experiences, provide actionable insights. Consistent with studies of health systems in fragile and conflict-affected settings, our results emphasize the importance of interdependence between sectors and proactive preparation to address unforeseen challenges.^26^ Moreover, our results reflect the cumulative impact of consecutive shocks, such as rapid Syrian and Yazidi resettlement programs and COVID-19, interact to compound health system demands over time.^26^ This compounding effect creates cascading burdens on health systems, demonstrating that such shocks cannot be effectively understood or managed in isolation.^27^

This study has limitations. The qualitative phase focused exclusively on health centre leadership, limiting insights into the experiences of other frontline staff and physicians. Additionally, we did not include refugee voices, critical to fully understanding the impact of health system shocks on care delivery, patient care perceptions, and outcomes. Finally, our analysis of COVID-19 only included its first nine months, precluding an assessment of long-term impacts on resilience and operations.

Despite these limitations, this study provides valuable insights into the resilience of specialized refugee health centres in Canada and how they respond to consecutive health system shocks. These findings may be generalizable to other refugee health settings in Canada, and practices and systems worldwide that provide care in the context of mass displacements. Further, this study illustrates that while specialized refugee beacon clinics can absorb, adapt, and transform in response to external shocks, sustaining these capacities requires adequate resources and comes with operational burdens, particularly to the well-being of frontline providers. As global geopolitical instability continues to drive displacement, proactive strategies to mitigate operational burdens will be critical to sustaining resilient refugee healthcare systems.

## Supporting information

Supplementary eTables

## Data Availability

Data Availability Statement: The data that support the findings of this study are available from the corresponding author upon reasonable request.

## Acknowledgements

We would like to thank the healthcare providers, staff and especially the patients at the Mosaic Refugee Health Clinic for their many contributions. Our sincere thanks for support from the Refugee Health YYC research team and the institutional support provided by the O’Brien Institute for Public Health, at the University of Calgary Cumming School of Medicine.

## Author Contributions

EN conducted data collection and analysis for both quantitative and qualitative phases, created visualizations, performed integration analysis, and led manuscript writing. LH double-coded qualitative data and contributed to integration and interpretation. GEF assisted in data analysis and contributed to interpretation of findings. RG improved the visual design of figures and diagrams. TW provided expertise in statistical methods and assisted in data analysis. LH, MYE, GEF, RG, KM, PR, RT, AC, and SM reviewed and edited the manuscript and approved the final version.

