## Supplementary eTables for "Refugee Healthcare Resilience and Burdens: A 10-year Mixed-Methods Analysis of System Shocks in Canada"

Supplement Exhibits

**eTable 1.** Level of Each Time Period and Significance of Difference Compared to Following

| Measurement | Baseline Initial (SD) | Baseline Final (SD) | Cuts Initial (SD) | Baseline vs. IFHP Cuts Level Comparison P-Value | Cuts Final (SD) | Syrian Surge Initial (SD) | Cuts vs. Syrian Surge P-Value | Syrian Surge Final (SD) | Yazidi Initial (SD) | Syrian Surge vs. Yazidi P-Value | Yazidi Final (SD) | COVID-19 Initial (SD) | Yazidi vs. COVID-19 P-Value | COVID-19 Final | Initial Baseline to Final COVID-19 P-Value |
|---|---|---|---|---|---|---|---|---|---|---|---|---|---|---|---|
| Mean Monthly Appointments for All Providers | 455.2 | 483.4 | 427.5 | 0.16 | 610.8 | 937.9 | **<0.01** | 1073.9 | 1179.9 | 0.29 | 1413.9 | 1214.4 | 0.11 | 2208.0 | **<0.01** |
| Mean Monthly Family Physician Appointments | 399.0 | 412.2 | 376.5 | 0.21 | 423.1 | 683.2 | **<0.01** | 614.9 | 627.2 | 0.86 | 562.7 | 719.3 | 0.12 | 1144.4 | **<0.01** |
| Mean Monthly Multidisciplinary Team Appointments | 31.9 | 37.1 | 26.2 | 0.48 | 156.5 | 216.3 | **0.01** | 404.1 | 493.3 | 0.11 | 628.4 | 618.8 | 0.88 | 1029.7 | **<0.01** |
| Mean Monthly Specialist Appointments | 33.9 | 32.6 | 23.7 | **0.02** | 33.9 | 39.8 | 0.14 | 54.3 | 59.4 | 0.21 | 66.3 | 32.9 | **<0.01** | 29.2 | 0.46 |

**Bold** denotes statistical significance ($p < 0.05$) in the difference between the level of the regression line at the last time point of one period and the first time point of the following period at a two-tailed alpha level of 0.05.

**Bold** denotes statistical significance ($p < 0.05$) in the difference between the initial baseline level and the final level at the end of the COVID-19 period at a two-tailed alpha level of 0.05.

**eTable 2**. Rate of Change in the Number of Monthly Appointments at a Specialized Refugee Health Centre

| Appointment Type | Baseline Rate* | IFHP Cuts Period Rate | Baseline vs. Cuts Rate of Change P-Value | Syrian Surge Rate | Cuts vs. Syrian Surge P-Value | Yazidi Period Rate | Syrian Surge vs. Yazidi P-Value | COVID-19 Period Rate | Yazidi vs. COVID-19 P-Value | Overall Rate of Change P-Value |
|---|---|---|---|---|---|---|---|---|---|---|
| All Providers | 1.6 | 4.5 | 0.45 | 9.1 | 0.48 | 6.3 | 0.78 | 110.4 | **<0.01** | **<0.01** |
| Family Physician | 0.7 | 1.1 | 0.89 | -4.5 | 0.32 | 2.5 | 0.29 | 64.6 | **<0.01** | **0.01** |
| Multidisciplinary Team (MDT) | 0.3 | 3.3 | 0.05 | 12.5 | **<0.01** | 3.7 | 0.1 | 45.7 | **<0.01** | **<0.01** |
| Specialists | 0.2 | 0.2 | 0.86 | 1.0 | 0.08 | 0.2 | 0.21 | 0.1 | 0.96 | 0.94 |

**Bold** values denote statistical significance in difference of slope ($p < 0.05$) at two-tailed alpha level of 0.05.
*Rate = change in appointments per month.

**eTable 3**. Provider Monthly Work Hours Supply

| Provider Type | Baseline Mean Monthly Hours (SD)* | Cuts Mean Monthly Hours (SD) | Syrian Surge Mean Monthly Hours (SD) | Yazidi Mean Monthly Hours (SD) | COVID-19 Mean Monthly Hours (SD) |
|---|---|---|---|---|---|
| All Providers | 415.9 (11.6) | 520.0 (94.6) | 975.3 (173.9) | 1392.7 (160.9) | 1649.3 (139.0) |
| Family Physicians | 320 (0.0) | 331.7 (42.2) | 554.7 (158.5) | 622.7 (118.0) | 736.0 (82.6) |
| MDT | 58.3 (12.7) | 150.9 (67.0) | 366.0 (48.5) | 691.5 (99.6) | 871.4 (89.8) |
| Specialist Physicians | 37.5 (4.8) | 37.4 (6.2) | 54.6 (8.0) | 78.6 (7.6) | 41.9 (9.5) |

*SD denotes standard deviation from the mean.

**eTable 4.** Good Reporting of A Mixed Methods Study (GRAMMS) Checklist

| Guideline | Section: page |
|---|---|
| Describe the justification for using a mixed methods approach to the research question | Introduction: Page 2. |
| Describe the design in terms of the purpose, priority and sequence of methods | Methods: Page 6-11 |
| Describe each method in terms of sampling, data collection and analysis | Methods: Page 6-11 |
| Describe where integration has occurred, how it has occurred and who has participated in it | Results: Page 14 |
| Describe any limitation of one method associated with the present of the other method | Discussion: Page 16 |
| Describe any insights gained from mixing or integrating methods | Discussion: Page 15-19 |
